# Quantification of SARS-CoV-2 viral copy number in saliva mouthwash samples using digital droplet PCR

**DOI:** 10.1101/2020.06.13.20130237

**Authors:** Lisa Oberding, Jia Hu, Byron Berenger, Abu Naser Mohon, Dylan R. Pillai

## Abstract

Saliva samples were collected through a simple mouth wash procedure and viral load quantified using a technology called digital droplet PCR. Data suggest ddPCR allows for precise quantification of viral load in clinical samples infected with SARS-CoV-2.

---

The shortage of nasopharyngeal (NP) swabs as sample collection devices has prompted clinical laboratories to explore other sample types. We conducted a community survey of patients previously diagnosed with COVID-19. Individuals were requested to gargle, swish, and spit 3 mL of normal saline (NS) into a standard sterile container from 12 individuals (REB 20-0444 and 20-0402, University of Calgary). Participants ranged from days 3 to 16 post symptom-onset. Four hundred μL of saliva in NS was subjected to RNA extraction using the Promega SV total RNA (Promega Corp., Madison, WI) following the manufacturer’s instructions; the initial lysis step was modified to add 400 μL of lysis buffer, wait 10 minutes, then add 400 μL 95% EtOH and proceed as normal. Two extracts were performed per sample. Extracted material was amplified using real-time PCR targeting the E gene with modification including the addition of GC clamps at the 3’ end of the primers and the shortening and addition of a minor groove binding (MGB) moiety to the hydrolysis probe (1). The sample extract was also evaluated by digital droplet PCR (QX200™ Droplet Digital™ (dd) PCR system, Bio-Rad laboratories, Hercules, CA) using the same primer set as used in the RT-PCR. Briefly, a master mix was made containing (per sample) 6.251⍰μL of One-Step RT–ddPCR Supermix, 2.51⍰μL of One-Step RT–ddPCR reverse transcriptase, 11⍰μL of 3001⍰mmol/L DTT, 11⍰μL of both forward and reverse E gene primers and 0.5 μL E gene probe (201⍰μM primers and 101⍰μM probe), 51⍰μL of RNA extract, and 7.51⍰μL of RNase-free water. Twenty microliters of each template mastermix was added to the sample well of the droplet generation cartridge, along with 70 μL of droplet generation oil for probes. Samples were cycled before measurement and analysis with the QX200™. The thermal cycling conditions were 50°C for 11⍰hour (reverse transcription) and 95°C for 101⍰min, followed by 40 cycles of 95°C for 301⍰sec and 60°C for 601⍰sec before a final step at 98°C for 10 min. Ramp rates were set to 2°C per sec.

Our data shows that the SARS-CoV-2 E gene is detectable in saliva obtained with a simple mouth wash approach using NS. The RT-PCR was able to detect the SARS-CoV-2 E gene in 9 of the 12 (75%) patient samples; however, ddPCR was able to detect copies of the E gene in 12/12 (100%) samples well above the no template control (NTC, n=14). The ddPCR data suggests that the majority of patients in this study had detectable viral gene copies in their saliva but the RT-PCR is only able to detect a subset of them. This is consistent with the exquisite analytical sensitivity obtained by ddPCR through partitioning the sample into 10-20,000 droplets from which PCR amplification of the template molecules occurs in each individual droplet. Furthermore, ddPCR enables the quantification of target nucleic acids at the single-molecule level. Depending on primer sets, the ddPCR system used in this study has a theoretical lower limit of detection between 2-4 copies per reaction (2). A recent study suggests that above a Ct value of 34 using the E gene RT-PCR assay sample are not infectious (3). Based on this information, 6/12 (50%) of saliva samples collected in this way are potentially infectious. Interestingly, patient 9 had a viral load estimated at 16,522 copies per 5 μL of extracted sample and E gene Ct value of 23.17 indicating high infectivity by all measures. These types of individuals may pose a risk in communal settings without isolation. We conclude that a simple mouth wash approach can be used to identify SARS-CoV-2 in saliva and that ddPCR may be useful in quantifying viral load with greater precision than reference RT-PCR.

**Figure 1.**
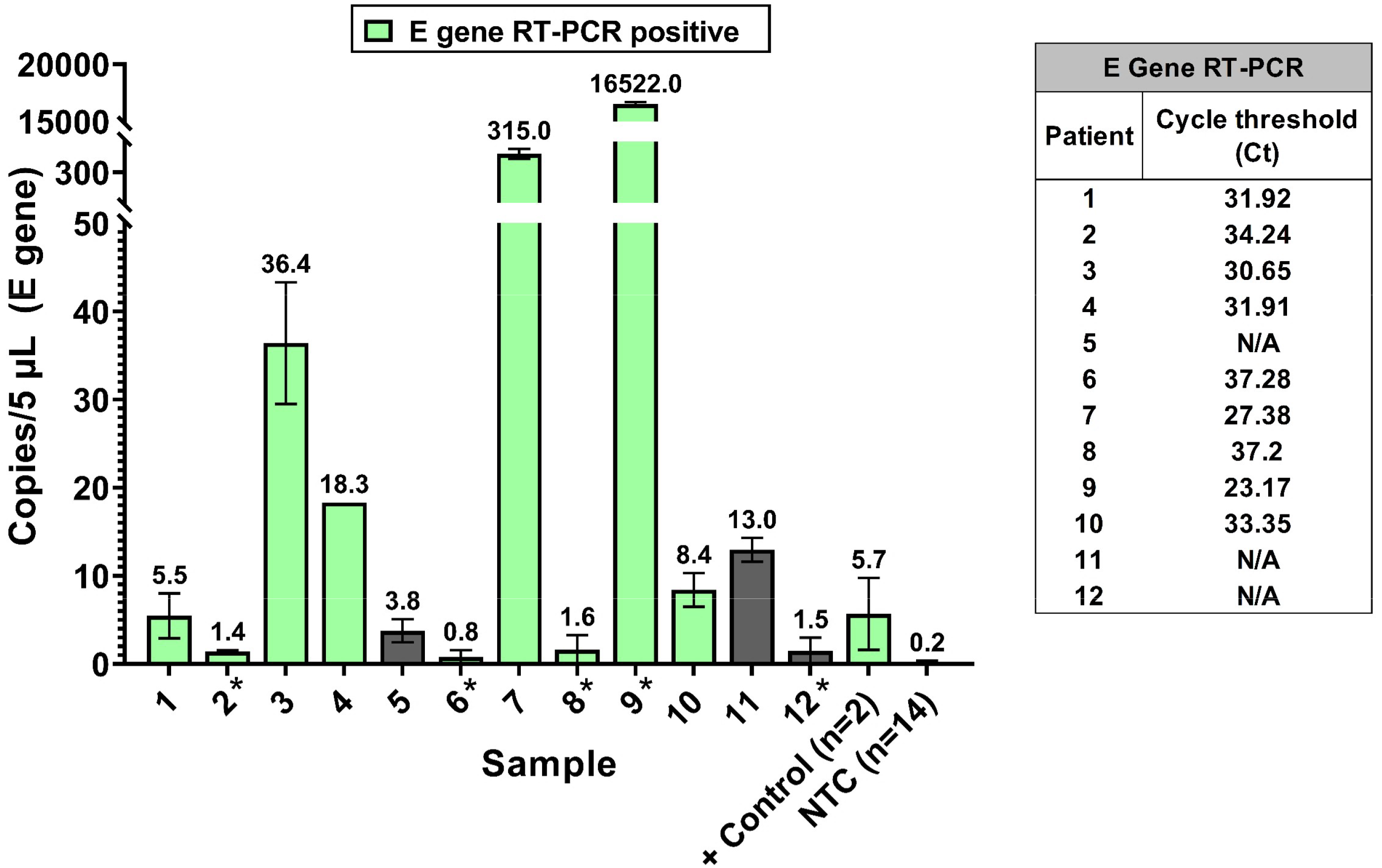
Correlation of E gene copy number and E gene RT-qPCR Ct value for saliva samples from patients infected with SARS-CoV-2. Graph shows ddPCR quantification (copies per 5 mL sample) with positive E gene RT-qPCR samples indicated in green. Copies are average of duplicate measurements with error bars showing SEM. Fourteen no template controls (NTC) were run. The ddPCR positive control contains 5 copies of template added per reaction. Ct values are shown below for E gene RT-qPCR for each corresponding sample. * Sample is either below or above the linear range of the QX200.

## Data Availability

Data is available upon reasonable request.

## Notes

### Competing Interest Statement

The authors have declared no competing interest.

### Funding Statement

The study was supported by funding from the Canadian Institutes for Health Research, Genome Canada, and Thistledown Foundation.

### Author Declarations

The study was approved by the IRB of the University of Calgary (CHREB REB20-0402 and REB20-044)

## References

1. Corman VM, Landt O, Kaiser M, Molenkamp R, Meijer A, Chu DKW, Bleicker T, Brünink S, Schneider J, Schmidt ML, Mulders Dgjc, Haagmans BL, van der Veer B, van den Brink S, Wijsman L, Goderski G, Romette J-L, Ellis J, Zambon M, Peiris M, Goossens H, Reusken C, Koopmans MPG, Drosten C. 2020. Detection of 2019 novel coronavirus (2019-nCoV) by real-time RT-PCR. Euro Surveill Bull Eur Sur Mal Transm Eur Commun Dis Bull 25.

2. Taylor SC, Carbonneau J, Shelton DN, Boivin G. 2015. Optimization of Droplet Digital PCR from RNA and DNA extracts with direct comparison to RT-qPCR: Clinical implications for quantification of Oseltamivir-resistant subpopulations. J Virol Methods 224:58–66.

3. La Scola B, Le Bideau M, Andreani J, Hoang VT, Grimaldier C, Colson P, Gautret P, Raoult D. 2020. Viral RNA load as determined by cell culture as a management tool for discharge of SARS-CoV-2 patients from infectious disease wards. Eur J Clin Microbiol Infect Dis Off Publ Eur Soc Clin Microbiol 39:1059–1061.

